# Protocol for update of food-based dietary guidelines for Czechia

**DOI:** 10.1101/2025.09.15.25335794

**Authors:** Eliška Selinger, Martin Krobot, Marina Jeřábková, Daniela Hrnčířová, Alexandra Košťálová, Eva Kupcová, Dana Kapitulčinová

**Affiliations:** Centre for Health promotion, National Institute of Public Health, Prague, Czech Republic; Department of Epidemiology and Biostatistics, 3rd Faculty of Medicine, Charles University, Prague, Czech Republic; Department of Public Health, Faculty of Medicine, Masaryk University, Brno, Czech Republic; Department of Hygiene, 3rd Faculty of Medicine, Charles University in Prague, Czech Republic; Environment Centre, Charles University, Prague, Czech Republic; Faculty of Humanities, Charles University, Prague, Czech Republic; Jan Evangelista Purkyně University in Ústí nad Labem, Ústí nad Labem, Czech Republic

## Abstract

**Background:** Current Czech national food-based dietary guidelines are outdated and do not reflect the most recent scientific evidence, nor considerations of sustainability and equity. Building on the Nordic Nutrition Recommendations 2023 (NNR), this project aims to develop updated, evidence-based food-based dietary guidelines (FBDGs) for the Czech Republic.

**Objectives:** To systematically update the evidence on associations between major food groups and health outcomes, integrate environmental and equity dimensions, and formulate clear, feasible, and context-specific recommendations through a structured consensus process.

**Methods:** For each food group, a PICO framework will be defined. Existing systematic reviews from NNR provide the evidence base; systematic “update” searches will cover the period from the NNR search cut-off until the present. Dual independent screening is performed, with artificial intelligence used as a secondary reviewer. Risk of bias will be assessed with RoB 2 (randomized trials), ROBINS-I (non-randomized interventions), ROBINS-E/RoB-NObs (observational exposure studies), and AMSTAR-2/ROBIS (systematic reviews). Certainty of evidence is graded with GRADE, complemented by NutriGrade where relevant. Parallel assessments summarize environmental impactsand equity/feasibility aspects. For each topic, concise evidence briefs will be prepared. Draft recommendation statements will be evaluated in a two-round modified Delphi process against five criteria (evidence consistency, clarity, feasibility/acceptability, equity, environmental impact). Consensus will be defined as a median score of 7–9 with less than one-third of ratings under 6.

**Discussion and dissemination:** The project will deliver transparently developed, evidence-based, and implementable food based dietary guidelines for the Czech Republic. Outputs will include final recommendations for the public and practice, a technical background report, and a record of comments and their resolution. All materials will be made openly available and presented to stakeholders, including public consultation groups.

## Introduction

Food-based dietary guidelines (FBDG) are a core public-health tool that translate nutrition science into practical, culturally appropriate advice for populations across the life course.^1^ They serve as a basis for food policy as well as educational activities and awareness rising. International guidance emphasises that FBDG should integrate not only health, but also dimensions of equity, and environmental sustainability, and be developed through transparent, evidence-based processes with explicit management of conflicts of interest (COI) and stakeholder input. In the European context, the 2023

Nordic Nutrition Recommendations (NNR)^3^ provide a comprehensive, methodologically advanced synthesis, including topic-specific systematic reviews and an explicit communication of environmental footprints, making them a suitable scientific basis for countries seeking to update national guidelines while adapting to local context.

Czech Republic, similarly to many Central and Eastern European countries, faces a high burden of non-communicable diseases (NCDs). Many of them are tightly related to known dietary risks and unhealthy diet is major contributor to the overall population burden of NCDs.^3^ Updating the national FBDG and their visual representation (e.g., plate/pyramid) therefore represents a timely public-health priority. However, guideline development in nutrition is challenging: much of the human evidence is observational, exposures are multidimensional and interlinked, and sustainability, affordability, and implementation constraints must be considered alongside known health effects. To ensure trust, reproducibility and policy relevance, methods must be prespecified, openly documented, and auditable.

Current Czech food-based dietary guidance relies on a food-pyramid tradition and textual recommendations that are now outdated: the Ministry of Health’s official FBDG date to 2005, later National Institute of Public Health (NIPH) materials on National Health Information Portal (NHIP) from 2013^4^ are used in practice but are themselves due for revision, and the prevailing pyramid has been criticised as insufficiently specific, didactically suboptimal, and missing broader nutrition issues (e.g., ultra-processing). At the same time, the diet-related disease burden is substantial: in 2017, an estimated 48% of deaths were attributable to lifestyle risks, with dietary risks alone accounting for one quarter pf premature death (23%), which is above the EU average. Adult obesity affected 20% of Czechs, with childhood overweight also increasing since the 1990s.^5^ These epidemiological data motivate a methodologically robust update that follows an established good practice, i.e., develop evidence-based textual FBDG first, with graphics (pyramid/plate) as the final step, and plan regular (∼5-year) revisions thereafter.

For these reasons, the update of FBDG was defined as one of the core tasks overview by NIPH in newly agreed National Cardiovascular Action Plan.^6^ The present protocol describes how the Czech FBDG (FBDG CZ) will be developed. It does not contain draft or final recommendations. Instead, it specifies (i) governance and COI management, (ii) evidence identification and appraisal, (iii) GRADE certainty assessment, (iv) integration of environmental sustainability and equity/feasibility evidence; and (v) consensus-building through a modified Delphi process. The protocol is based on established methodological standards for evidence synthesis, risk-of-bias appraisal, and guideline development, adapted to the specific needs of food-based guidance and the Czech implementation setting. Simplified version of the protocol will also be submitted to a local professional journal to ensure local community has access to relevant information in native language.

### Brief overview of the process

NNR offers state-of-the-art systematic reviews and certainty ratings across major food groups and dietary patterns, along with explicit sustainability considerations. Building on this resource increases efficiency and avoids duplication, especially in the context of limited public health system capacities. It also improves comparability across European contexts. For topics with sufficient NNR coverage, we will conduct systematic updates from the last NNR search date. For topics insufficiently covered or evolving rapidly, such as plant-based dairy substitutes or alternative proteins, we will conduct de novo systematic reviews with full protocolised methods. In both cases, we will reassess risk of bias with current tools and rate certainty using GRADE.

Environmental impacts^7^ will be calculated for the current Czech dietary pattern and for the proposed health-optimised scenario, using the best available life cycle assessment (LCA) data of food products from the closest regional context. Where Czech-specific data are unavailable, regional datasets most representative of Czech supply chains and consumption patterns will be selected. Changes between the two situations will be assessed to guide final recommendations with respect to environmental sustainability. Equity will be addressed considering socioeconomic status, geography, age, disability, and other, with the focus on affordability, availability, culinary skills/time, and cultural acceptability. These inputs will later guide the wording of public-facing recommendations.

Formulating concise, actionable wording requires structured expert judgement based on overall consensus. We will use a two-round modified RAND/UCLA Delphi^8^ with multidisciplinary panelists, representing wide range of stakeholders, professional representatives, as well as members of general public, but excluding individuals with unmanaged COI. Panel members will rate candidate sentences on evidence alignment, clarity, feasibility/acceptability, equity impact and environmental alignment using 1–9 scales, with predefined consensus rules. Stakeholders, including industry, may submit written comments during preparation of background materials, all managed in a transparent consultation process but will not set PICO questions, assess or grade evidence, or vote on final wording. All COI will be declared on WHO Declaration of Interests (DOI) forms.

Nutrition guidelines shape consumer demand, public procurement, school meals, labelling and fiscal policies. As such, they are targets for organised influence by commercial actors with direct financial stakes in recommended (or discouraged) products.^9^ A large body of empirical research shows that industry sponsorship is associated with more favourable conclusions and selective framing in nutrition and health research, through design choices, selective outcome reporting, or interpretation biases, raising the risk of distorted evidence bases if safeguards against such influence are weak.^10^ Guidance from WHO and leading methods organisations therefore emphasises structural independence, mandatory, standardised disclosure of financial interests, prospective management plans or stakeholder engagement that separates consultation from evidence appraisal and voting.^11–13^ In line with these standards, our protocol (i) requires WHO based DOI for all contributors, including annual updates; (ii) excludes current employees and office-holders of food, beverages, supplement or alcohol companies and their trade associations from decision-making bodies, and (iii) require industry participation only via transparent written submissions during public consultation with stakeholder groups. These measures are necessary to preserve trustworthiness and legitimacy, protect against systematic bias in evidence interpretation, and ensure recommendations reflect public-health goals rather than commercial priorities.

### Objectives

1. To pre-specify the governance, COI management, and transparency mechanisms for FBDG CZ.
2. To define topic-specific PICO frameworks, based on NNR where applicable
3. To detail study identification, selection, data extraction, and risk-of-bias procedures
4. To outline GRADE certainty assessment and integration of sustainability and equity/feasibility evidence into decision-making.
5. To describe the Delphi consensus and public consultation processes, including their documentation

By publishing this protocol on an open repository, we seek to improve credibility, reusability, and policy uptake of the resulting Czech FBDG, while enabling external scrutiny and future updates.

## Governance and Conflict-of-Interest (COI) Management

### Organisational structure and roles

The FBDG CZ will be developed utilising a partnership between academia and public sector, with a pre-specified independence policy.

We will establish three main groups of bodies with distinct mandates:

- **Technical Working Group (TWG)**. Multidisciplinary experts in epidemiology, nutrition, public health, or environmental assessment. The TWG is responsible for scoping and defining PICO per topic, approving search strategies, risk-of-bias (RoB) and GRADE judgements, synthesis of identified evidence, assessing environmental impacts, and drafting candidate public-facing statements. Members must meet eligibility and COI criteria (see below). The TWG is lead by NIPH, with members invited from leading Czech academic institutions.
- **Consultative Stakeholder Groups (CSGs)**. Open, non-voting forum including education and catering sectors, civil society, professional (medical) societies, and the agri-food industry. The CSGs can submit written comments and data during public consultations and targeted calls,especially on selection of topics and during evidence review and synthesis; they do not set PICO, appraise evidence, or vote on draft or final recommendations, consistent with guidance on conflict of interest management and independence in guideline processes.
- **Advisory Panel:** Representatives of experts in nutrition, education, public health, communication as well as members of public, who will be involved in the Delphi process. Members will be invited based on their professional profile or external nominations and should represent experts from Czechia as well as abroad, where relevant. All members will be required to meet COI criteria confirmed by COI form to ensure protection from commercial interference.

### Membership eligibility and decision-making

All TWG and Advisory Panel members will complete WHO DOI forms at entry and annually thereafter and undergo COI screening before assignment to any topic. TWG members will also complete onboarding on TWG procedures, GRADE, RoB tools, and use of the Evidence-to-Decision (EtD) framework. Selection prioritises methodological competence, topic expertise, absence of relevant COI, and diversity across disciplines and perspectives. A list of TWG members will be made publicly available, with members of the Advisory Panel published after closure of Delphi process.

TWG decisions on PICO definitions, RoB judgements, GRADE ratings, and candidate statements require a quorum (≥ two-thirds of voting members) and simple majority voting. Where votes are split, minority positions will be documented.

### COI policy: declaration, assessment, and management

We adopt a COI policy that is inspired by WHO’s Framework of Engagement with Non-State Actors (FENSA)^13^, WHO DOI for experts, and overall approaches described in the WHO “*Draft approach for the prevention and management of conflicts of interest in the policy development and implementation of nutrition programmes at country level*.”^14^

All financial interests in the previous 36 months (employment, advisory roles, speaking fees, consultancy, research funding, stock/stock options, patents/royalties, paid expert testimony) and relevant non-financial interests (e.g. unpaid leadership in relevant organizations), plus any interests held by close family members where relevant. Reported COI will be categorized by topic relevance (general vs topic-specific), magnitude (minor/moderate/major), and recency. Determinations and rationales resulting from the TWG discussion will be recorded.

Depending on risk, actions include disclosure only, speaking but not leading, recusal from specific agenda items, recusal from all decisions on a topic, or ineligibility for TWG or Advisory Panel service. Individuals with current employment by, or governance roles in, companies/trade associations whose primary business is the manufacture or marketing of foods, beverages, supplements or alcohol or related inputs are ineligible for TWG and Advisory Panel membership. They may participate only via the CSGs written-comment process. Topic leads cannot have topic-specific financial COI.

### Stakeholder engagement boundaries and consultation

Stakeholder input improves feasibility and legitimacy but may introduce bias if the conflicting interests are not managed well. Consistent with available guidance, stakeholders (including industry) may provide written evidence and implementation data, comment on drafts during consultations, and report any potential unintended consequences or equity concerns. However, they may not set PICO questions, screen/extract/appraise evidence, perform or influence GRADE ratings or vote on recommendation wording. All submissions will be logged and made public with dispositions (accept/partial/reject) and rationale for the decisions.

### The consensus processes

The TWG prepares evidence briefs and candidate statements, while a modified RAND/UCLA Delphi panel (separate from CSG and industry) rates statements on five criteria (evidence alignment, clarity, feasibility/acceptability, equity impact, environmental alignment). Consensus thresholds and documentation standards are prespecified. DOIs and recusals for Delphi panelists follow the same procedures as for TWG.

## Evidence review and synthesis

### Design overview

This is a methods protocol for an umbrella review based on NNR2023 topic-specific systematic update reviews combined with de novo systematic reviews for topics insufficiently covered or newly emergent. Methods follow PRISMA guidelines for search reporting, with risk-of-bias (RoB) tools appropriate to study design, GRADE for certainty of evidence, and an explicit plan to integrate environmental sustainability and equity/feasibility into evidence-to-decision considerations. No recommendations are formulated in this protocol.

### Registration and search strategy

Protocol is published on an open pre-print server. Where feasible and needed, topic-level review records will be registered (e.g., PROSPERO for eligible de novo reviews). For topics based directly on NNR, searches will update from the last NNR search date through 2025-08-31. Additional search will be repeatedly conducted before the initiation of the Delphi process to ensure no relevant recent evidence will be omitted. For de novo topics, searches cover inception (or an agreed historical start date, if justified) through 2025-08-31.

Topic-specific strategies will combine controlled vocabulary and text words for exposure/food group, outcomes, and study design filters, adapting NNR terms where relevant. Searches are limited to human studies with no language restrictions at the search stage. Reviewers will screen titles/abstracts and full texts. To increase the efficiency of the screening process, the screening of titles and abstract will be conducted by human researchers in parallel with AI, as a second control. Disagreements will be resolved by consensus in TWG. Reasons for exclusion at full text will be recorded. A PRISMA 2020 flow diagram will be produced for each topic.^15^

### Eligibility criteria

#### Population

General populations or at-risk groups relevant to primary prevention across the life course (including pregnancy, lactation, children, older adults).

#### Exposures

Food groups, dietary patterns, or processing levels (e.g., NOVA) with clearly defined units (g/day, servings/week) or categorical contrasts. Nutrient-only exposures are excluded unless mechanistically essential for a food-based question.

#### Comparators

Lower or no intake, alternative food(s), or usual care;

#### Outcomes

- **Primary:** All-cause mortality; cardiovascular disease (CVD) incidence/mortality; type 2 diabetes (T2D); site-specific cancers; adiposity/weight change.
- **Secondary:** blood pressure, lipids, glycaemia, kidney outcomes, bone health, micronutrient adequacy, oral health, pregnancy/infant outcomes where applicable; **harms** (e.g., contaminants, allergy, food safety).

#### Study designs

Existing systematic reviews will be used as a main basis for FBDG CZ, combined with individual observation or intervention studies when deemed necessary. In that case, minimum follow-up of ≥1 year for incident NCD/mortality in observational cohorts will be required, with shorter durations permitted for biomarkers (secondary outcomes) and trials.

#### Exclusion criteria

Ecological studies, narrative reviews, case series, and disease-management

### Risk of bias (RoB) appraisal

RoB will be assessed in duplicate using design-appropriate standard tools; disagreements will be resolved by the consensus of the TWG. Funding source and author COI will be recorded as potential bias.

For systematic reviews, which should serve as a basis for the FBDG CZ, AMSTAR-2^16^ will be used to assess the methodological quality and ROBIS^17^ to assess risk of bias. In case of the need to evaluate primary studies, the observational studies of exposures (e.g., cohorts on foods/patterns) will be assessed using ROBINS-E^18^/RoB-NObs tool. For assessment of randomised trials (including cluster or crossover), RoB 2^19^ will be selected as a tool, while non-randomised intervention studies (quasi-experiments) will be assessed by ROBINS-I.^20^

### Evidence synthesis

For health related outcomes, primary effect measures will be risk ratios (RR) or hazard ratios (HR) for incidence/mortality and mean differences (MD) or standardised mean differences (SMD) for continuous outcomes. For each included study, we will extract funding sources and author COI disclosures to inform overall certainty in evidence evaluation and overall synthesis.

The certainty of evidence will be rated per outcome and comparison using GRADE^21^ accompanied by NutriGRADE^22^ with explicit rationales.

GRADE will be used as the primary, standardised framework for rating the certainty of evidence per outcome and comparison. It is the international reference used in guideline development and aligns with our Evidence-to-Decision approach. Under GRADE, randomised trials start at High certainty of evidence and observational studies at Low certainty, ratings are then downgraded for concerns in domains of risk of bias, inconsistency, indirectness, imprecision, and publication bias, or upgraded when there is a large effect, a clear dose–response gradient, or when all plausible residual confounding would reduce (rather than increase) the observed effect. Final levels of certainty and their interpretation are described in Table 1. However, despite being a standard tool for guideline development, GRADE is not without criticism, especially in nutrition. Its critiques point out that GRADE systematically downgrades bodies of evidence from topics in which usage of randomized controlled trials is limited for practical or ethical reasons and for which we need to rely on prospective cohorts. To increase transparency and nutrition-specific conclusions, we will therefore assess the evidence with parallely with NutriGRADE. NutriGRADE sums eight items, including study limitations/risk of bias, precision, heterogeneity, directness, publication/funding bias and reports categorical cut-points. The comparison between GRADE and NutriGRADE is summarized in Table 2. GRADE remains the main base for judgments and decisions; NutriGRADE is used only as a supplementary nutrition-specific check, and any divergences will be explicitly explained.

**Table 1:** GRADE assessment of certainty of evidence interpretation.

| GRADE: Certainty of evidence rating |  |
| --- | --- |
| High certainty (⊕⊕⊕⊕) | • Further research is very unlikely to change the confidence in the estimate.<br>• Usually: for nutrition questions where RCTs are impractical, a large, consistent body of prospective observational evidence that satisfies GRADE upgrade criteria (large effect, dose–response, and plausible residual confounding would attenuate the effect) and has no downgrades may also qualify. |
| Moderate certainty (⊕⊕⊕○) | • Further research is likely to have an important impact on the confidence and may change the estimate.<br>• Usually: downgraded one level for a single serious limitation in any domain below. |
| Low certainty (⊕⊕○○) | • Further research is very likely to have an important impact and likely to change the estimate.<br>• Usually: downgraded two levels (e.g., two serious or one very serious limitation); or observational evidence that has been upgraded once. |
| Very low certainty (⊕○○○) | • There is a very little confidence in the effect estimate; the true effect is likely to be substantially different.<br>• Usually: severely downgraded bodies of evidence; or observational evidence with serious issues and no upgrade factors. |

**Table 2:** Comparison of GRADE and NutriGRADE assessment.

| Points | Domain | GRADE | NutriGRADE |
| --- | --- | --- | --- |
| Downgrading points | Risk of bias | Downgrades if trials/cohorts have important limitations | Scores study limitations of included studies; higher points for low risk of bias across studies; fewer points if major concerns are common. |
|  | Inconsistency | Downgrades for substantial, unexplained heterogeneity (e.g., widely varying effects, high I<sup>2</sup>, non-overlapping CIs). | Points reduced when there's notable, unexplained heterogeneity; full points when estimates are consistent |
|  | Indirectness | Downgrades if population, exposure, comparator, or outcomes don't directly match the question; surrogate endpoints | Penalizes indirectness (e.g., surrogate markers, non-target population/food matrices); full points for direct evidence |
|  | Imprecision | Downgrades for wide CIs that include both benefit and harm or fail to meet optimal information size | Awards points for adequate precision and CIs not spanning key decision thresholds; fewer points if imprecise |
|  | Publication bias | Downgrades if small-study effects or missing negative studies are suspected | Deducts points when funnel plots/tests suggest bias or when evidence base is small/asymmetric |
|  | Funding bias/conflict | Considered within risk of bias and publication bias; not a separate, explicit domain | Explicit item: points deducted if industry funding/sponsorship likely biases results or reporting |
| Upgrading points | Large magnitude of effect | Upgrade observational evidence for large or very large effects (no obvious bias) | Explicit scoring: more points for larger, credible effects |
|  | Dose-response gradient | Upgrade if a clear dose-response is shown | Explicit scoring: points for consistent dose-response across exposure categories |
|  | All plausible confounding | Upgrade if confounding would reduce (not explain) the observed effect | Reflected indirectly via study quality/funding bias and effect credibility; not a separate scored item |

Moreover, the credibility of evidence will be evaluated, using pre-specified evidence classification criteria and categorized as convincing (“class I”), highly suggestive (“class II”), suggestive (“class III”), weak (“class IV”) or no evidence (“class V”; Table 3)^23^. Summary tables will provide measures of effect, risk of bias notes and certainty and evidence classification ratings (high/moderate/low/very low).

**Table 3:** Criteria for classification of credibility of evidence.

| The evidence classification criteria for the credibility of evidence* |  |
| --- | --- |
| Convincing (Class I) | • The number of cases is $> 1000$ (or $> 20,000$ for continuous outcomes)<br>• $P < 1 \times 10^{-6}$<br>• $I^2 < 50\%$<br>• 95% prediction interval excludes the null hypothesis<br>• No small-study effects<br>• No excess significance bias |
| Highly suggestive (Class II) | • Class I criteria not all met<br>• The number of cases is $> 1000$<br>• $P < 1 \times 10^{-6}$<br>• The largest included individual study exhibits $P \leq 0.05$ |
| Suggestive (Class III) | • Class I–II criteria not all met<br>• The number of cases is $> 1000$<br>• $P < 1 \times 10^{-3}$ |
| Weak (Class IV) | • Class I–III criteria not all met<br>• $P \leq 0.05$ |

\* Adapted from Lane M M, Gamage E, Du S, Ashtree D N, McGuinness A J, Gauci S et al. Ultra-processed food exposure and adverse health outcomes: umbrella review of epidemiological meta-analyses *BMJ* 2024; 384 :e077310 doi:10.1136/bmj-2023-077310

For each food group, evidence on relation to greenhouse-gas emissions, land and water use, eutrophication, and biodiversity impacts will be assessed and reported. Similarly, evidence on availability, affordability (price per serving), cultural acceptability, culinary skills/time, and constraints in school/public catering will be gathered and discussed. These inputs will inform EtD considerations and help anticipate differential effects across population subgroups.

All acquired evidence and relevant inputs for individual food groups will be narratively summarized to a report, serving as a basis for formulation of public-facing statements finalized during the following Delphi processes.

### Stakeholder consultation, public comments, scope and safeguards

Public consultation is a core component of trustworthy guideline development. It can improve clarity, address contextual constraints (affordability, availability, culture), and strengthen legitimacy and uptake among those who will implement and live with the guidance. International standards explicitly recommend transparent, time-bounded consultation with broad participation (public, patients, practitioners, service providers)^11^ to reduce blind spots and to incorporate diverse values and preferences in EtD considerations, but without reopening evidence appraisal itself. If performed correctly, public consultation supports equity by giving voice to under-represented groups (e.g., lower-income households, schools/public catering).

At the same time, consultation is highly sensitive to commercial determinants of health^24,25^ and other organised influences: astroturfing and orchestrated volume campaigns, selective citation of low-quality evidence, front-group submissions, and reputational pressure. All of these can bias processes if safeguarding rules are weak. The food and alcohol sectors have well documented corporate political activities (framing, coalition management, information and financial strategies) that can seriously distort nutrition policy debates and put public health at risk.^9,26,27^ To ensure that the interests of public health are treated as a main priority during the development of FBDG CZ, which we view as a core health-oriented tool, we will adopt approach consistent with WHO approach and COI guidance, with management focused on well-described commercial strategies. Our process will therefore (i) separate consultation from PICO setting, evidence appraisal and voting, (ii) require name/affiliation and DOI for submissions of comments, (iii) aggregate form-letter campaigns as a single issue in the comment matrix, (iv) weight responses by argument merit and evidentiary quality, not by count, and (v) publish a full comment matrix with dispositions and rationales. These safeguards maximise the benefits of open input and transparent public debate, while protecting the guideline’s independence, credibility, and public-health focus.

## Summary of public consultation process

### Scope

Evidence synthesis including appraisal (RoB/GRADE) will be distributed among members of the CSGs and made publicly available. Comments submitted via special online form or through written communication at CSGs will be welcomed and assessed by the TWG.

### Who can comment

public, professional societies, patient groups, NGOs, education sector, catering providers, and industry. Submissions for comments will require name/affiliation and COI disclosure.

### Boundaries

CSGs and public will not set PICO, screen/extract/appraise evidence, or vote on statements. Engagement will follow our COI policy. Public consultation will provide non-voting input primarily on clarity, feasibility/acceptability and equity. Evidence appraisal and evidence-alignment ratings will be determined through the expert Delphi process and are not subject to public voting.

### Process

Submissions via web form, will require name/affiliation; comment; proposed edit; evidence supporting the need for edit and declaration of COI. The submission of comments will be open for 2-4 weeks.

### Disposition

A Comment Matrix will record each submission with disposition (accept/partial/reject) and rationale.

### Deviations and amendments

Any deviation from this protocol (e.g., modification of eligibility, additional analyses) will be prospectively recorded in an Amendments Log with date, rationale, and implications for interpretation.

## Delphi Consensus and Stakeholder Consultations

### Objectives

To convert developed evidence syntheses into clear, feasible, equitable, and environmentally aligned public-facing statements via a modified RAND/UCLA Delphi (two rounds)^8^, and to invite broader input through a public consultation that will not interfere with evidence appraisal or voting. Detailed protocol for the Delphi process will be developed. However, a brief outline of the proposed process is described below.

### Panel composition, eligibility, and recruitment

The planned target for panel size is 20-40 multidisciplinary experts (nutrition, epidemiology, medicine, public health practice, food hygiene, environmental assessment, behavioural/implementation science, health economics, school/public catering). Delphi sample sizes are not power-based, but planned range represents a usual practice which balances diversity with operational feasibility. Invitations will include role description, COI policy, and expected time per round. Replacement of non-responders is not planned after the start of Round 1 to avoid bias.

All panelists will complete WHO DOI at entry, with topic-specific COI screening and recusal rules described in the previous section. Individuals employed by or governing food, beverages, supplement or alcohol companies or trade associations are ineligible. Industry stakeholders may comment only via the public consultation processes.

### Materials provided to panelists

For each topic (food group), panelists receive a standard package consisting of:

- **Evidence Brief** (1–2 pages long; health effects with key pooled estimates, RoB and GRADE).
- **Sustainability and equity/feasibility Brief** (1-2 pages long)
- **Candidate statements** (1–7 per topic), each with one-line rationale

**Delphi processes criteria and management**

For the Delphi process, panelists will be asked to evaluate every statement provided on 5 dimensions. For every dimension, 9-point Likert scales will be used as recommended in RAND/UCLA:

A. **Evidence alignment** (1 = contradicts evidence; 9 = fully aligned with evidence)
B. **Clarity/unambiguity** (1 = unclear; 9 = crystal-clear)
C. **Feasibility/acceptability** in CZ context (1 = impractical/unacceptable; 9 = readily feasible)
D. **Equity impact** (1 = likely worsens inequities; 9 = likely improves inequities)
E. **Environmental alignment** (1 = misaligned with sustainability; 9 = well aligned with sustainability)

Free-text comments will be required whenever a rating ≤6 is given on any criterion. Evidence and environmental alignment will be evaluated solely by the topic experts in the panel, while the rest of dimensions will be evaluated by the full panel. Ratings will be anonymous to peers, coordinators will be able to link identities to DOIs and recusals. The answers will be gathered through web survey and GDPR-compliant data handling will be employed. A short guideline will be provided to panelists, including exemplar rating exercise, which should ensure shared understanding. Round 1 will be open for 28 days (reminders on days 7, 14 and 25); Round 2 will be open for 14 days.

For each statement × criterion median and interquartile range (IQR) will be computed, together with the RAND Disagreement Index (DI). The group medians/IQRs and anonymised comment themes will be circulated among all panelists. After the first round, the TWG will revise statements focusing on clarity and feasibility (evidence re-appraisal only if clearly warranted), adapt the statements based on feedback provided, and prepare Round 2 packages. During the second round, the revised statements will be re-rated. In case that DI remains high with no pathway to clarity, the statement can be dropped, minority report recorded or in rare cases Round 3 initiated, based on the decision of TWG.

### Consensus definition and decision rules

A statement achieves consensus when, for each criterion:

- Median is 7–9, and
- ≤ one-third of ratings are ≤6, and
- DI < 1.0 (RAND criterion) indicating no substantive disagreement

Statements failing one or more criteria are revised or discarded with rationale. The final set will be the Delphi-endorsed wording of FBDG CZ forwarded to the design/communication phase (outside this protocol).

### Panel safeguards, criterion-specific voting, and rating validity

To preserve scientific integrity while ensuring meaningful public and patient input, several steps will be employed during the process: stratification of voting by criterion, guidance for panelists and enforcement of rating validity rules.

### Criterion-specific voting

For A. Evidence alignment and E. Environmental alignment, only subject-matter and methods experts provide ratings, all panelists, including patient/general-public representatives, rate B. Clarity, C. Feasibility/Acceptability, and D. Equity.

### Guidance

Before Round 1, all panelists will receive a guidance and sign an attestation that ratings for criterion A will be grounded in the provided evidence, while personal preferences will be recorded in free-text comments.

### Validity of ratings

Any rating ≤6 requires a justification. For criterion A and E, justifications must reference specific evidence supporting the comment. Ratings without such justification are coded procedurally invalid (NA) for criterion A and E and excluded from consensus evaluation.

### Consensus rules

A statement reaches consensus when, for each criterion, the median is 7–9 and ≤1/3 of valid ratings are ≤6. The expert-only median for A is the decisive metric, with all-panel medians shown for transparency. Disagreement will be quantified using the RAND Disagreement Index.

### Recusals

Topic-level recusals may be invoked when a panelist’s declared interests that would reasonably compromise judgement. All recusals are logged and reported.

### Data protection and ethics

The Delphi process involves expert opinion with minimal risk. Participation is consent-based and data stored in compliance with GDPR (EU 2016/679).^28^

### imitations and mitigation

Consensus methods may over-smooth dissent or bias towards the status quo. We plan to mitigate this via anonymity, explicit disagreement diagnostics (DI), documentation of minority positions, and strict COI/recusal enforcement. Public consultation can introduce volume-based biases, a structured Comment Matrix and transparent dispositions reduce undue influence.

## Conclusion

This protocol specifies a transparent workflow and rationale for updating the Czech food-based dietary guidelines. By anchoring topic work in Nordic Nutrition Recommendations 2023 and conducting systematic updates (or de novo reviews where needed), applying design-appropriate risk-of-bias tools, and grading certainty and credibility of evidence, we aim to generate an evidence base that is scientifically credible and with transparent evidence to decision processes, with safeguarding of independence through rigorous COI management and stakeholder and public engagement. A modified RAND/UCLA Delphi will translate evidence into concise, implementable public-facing statements, while public consultation will improve clarity and feasibility without compromising methodological and scientific integrity.

All produced reports will be publicly available. Together, these methods are designed to produce trustworthy, context-appropriate guidance that can be implemented across households, schools, healthcare, and public procurement, and that can be periodically revised as new evidence and monitoring data emerge.

### Next steps: development of graphical representation

Following finalization of the textual FBDG wording (post-Delphi), we will commission and develop its graphical representation (e.g., plate/pyramid or equivalent schema). Visuals will not introduce new targets and they will strictly reflect the endorsed recommendations. The design process will include concept generation as well as user testing with diverse subgroups (adolescents, older adults, lower-income households, and food-service professionals) and including development of ready-to-use materials for schools or public catering. The final graphical representation will be released and updated in tandem with future guideline revisions.

## Data Availability

No data were produced.

